# Alzheimer’s Disease Classification Using Cluster-based Labelling for Graph Neural Network on Heterogeneous Data

**DOI:** 10.1101/2022.03.03.22271873

**Authors:** Niamh McCombe, Jake Bamrah, Jose M. Sanchez-Bornot, David P. Finn, Paula L. McClean, KongFatt Wong-Lin, Alzheimer’s Disease Neuroimaging Initiative (ADNI)

**Author notes:** Correspondence: KongFatt Wong-Lin. Joint first authors (Niamh McCombe, Jake Bamrah).

## Abstract

Biomarkers for Alzheimer’s disease (AD) diagnosis do not always correlate reliably with cognitive symptoms, making clinical diagnosis inconsistent. In this study, the performance of a graphical neural network (GNN) classifier based on data-driven diagnostic classes from unsupervised clustering on heterogeneous data, is compared to the performance of a classifier using clinician diagnosis as outcome. Unsupervised clustering on tau-PET and cognitive and functional assessment data was performed. Five clusters embedded in a nonlinear UMAP space were identified. The individual clusters revealed specific feature characteristics with respect to clinical diagnosis of AD, gender, family history, age, and underlying neurological risk factors. In particular, one cluster comprised mainly diagnosed AD cases. All cases within this cluster were re-labelled AD cases. The re-labelled cases are characterised by high cerebrospinal fluid amyloid beta (CSF Aβ) levels at a younger age, even though Aβ data was not used for clustering. A GNN model was trained using the relabelled data with a multiclass area-under-the-curve (AUC) of 95.2%, higher than the AUC of a GNN trained on clinician diagnosis (91.7%; p=0.02). Overall, our work suggests that more objective cluster-based diagnostic labels combined with GNN classification may have value in clinical risk stratification and diagnosis of AD.

## 1 Introduction

Alzheimer’s disease (AD) is the most common form of dementia and is commonly manifested through a variety of symptoms such as cognitive degradation, motor impairment, speech disturbance and psychiatric changes (1). As of 2019, between 60-70% of all dementia cases are AD cases, with the disease predominantly affecting those aged 65 and over [1]. Clinical diagnosis of AD is not always consistent, partially due to inconsistent correlation of AD disease stage with known AD biomarkers [2-4].

With the availability of increasingly complex and heterogeneous dementia data and still sub-optimal dementia diagnostic procedures, decision support systems, with the aid of machine learning (ML) algorithms, are gradually becoming important [3]. However, most ML classification on AD data makes use of clinician diagnosis to supervise the learning. There has been no utilization of more objective datadriven labelling of classes using unsupervised ML. Particularly, although previous studies demonstrated benefits of unsupervised clustering in identifying sub-groups of patients (e.g., [5-7], none of the studies has used the identified clusters for AD classification.

In terms of ML for classification, there has been an increase in a specific approach – graphical neural networks (GNNs) [8]. A variety of GNN approaches have been applied on different types of data ([8-10]. A limitation of such GNN based approaches was that of graph rigidity, where the final graph structure was limited when inducting new nodes. Such an example was provided in [9] where the resulting model required a new model iteration to be trained with each new node addition. More robust GNN techniques have been proposed ([11, 12]). In particular, in [12], a flexible GNN was proposed to solve the fixed graph structure problem by adopting a meta-learning strategy, specifically metric-learning, which was used to infer node similarity using a trainable similarity function, facilitating the use of heterogeneous data types, including magnetic resonance imaging (MRI) data.

GNN studies, to date, have not made use of tau-specific positron emission tomography (PET) neuroimaging data, one of the key lesions in AD [3]. Studies have suggested that tau PET brain images readily matched the distribution of tau deposits reported from histopathological studies, brain atrophy, hypometabolism and overall severity of AD [13] and may be better than cerebrospinal fluid amyloid-beta (CSF Aβ), amyloid-PET and MRI in AD prognosis (e.g. [14, 15]). In this study, we address the above limitations by first applying nonlinear dimensional reduction on a heterogeneous dataset, which includes tau PET neuroimaging data coregistered with MRI, and individual sub-assessments from cognitive and functional assessments. This is followed by data clustering, and then we investigate the feature characteristics of the individual clusters. Next, we re-label cases based on cluster information, validated by tau-PET data and data on Aβ, which was not used for clustering, to form new classes of AD and non-AD for GNN’s AD classification. Finally, the GNN’s performance using the re-labelled data is compared with that using clinician diagnosis.

## 2 Methods

### 2.1 Data Description

The dataset used in this study was obtained from the open Alzheimer’s Disease Neuroimaging Initiative (ADNI) database (adni.loni.usc.edu), particularly the ADNIMERGE3 open repository. A complete workflow of data preparation, processing and analysis is shown in Figure 1. Only ADNI participants who had undergone MRI and tau PET scans (for detecting tau deposition) were extracted from the data, and the PET and MRI data were merged with sociodemographic, medical/family history and neuropsychological features as measured at study baseline. The final dataset comprised 224 features: 7 sociodemographic and medical history features, 40 cognitive and functional assessments’ (CFAs) scores, and 177 neuroimaging features (from combined MRI and tau PET imaging data; see below). Clinician diagnosis of participants, considered as a class label for training GNN model (see below) consisted of control normal (CN), Alzheimer’s Disease (AD), and mild cognitive impairment (MCI – which includes prodromal stage of AD).

**Figure 1.**
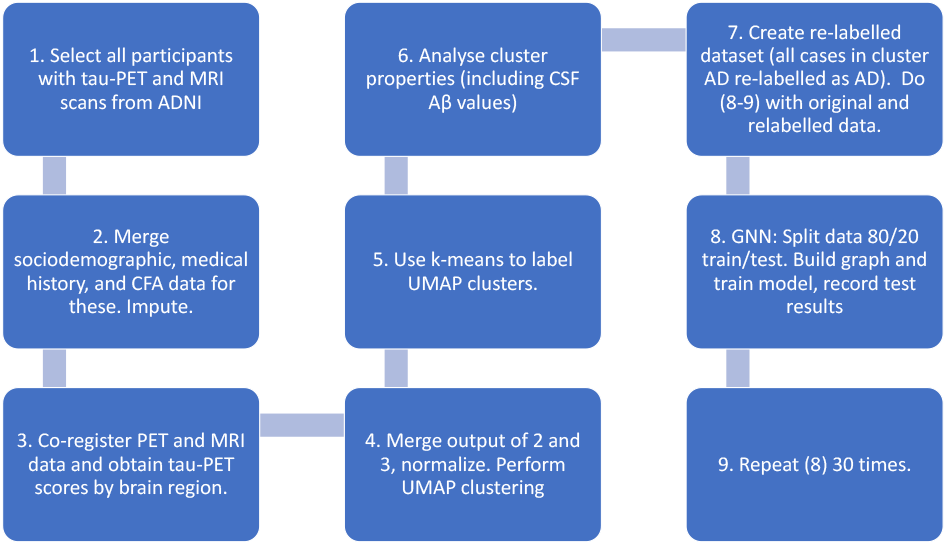
Data processing and analytical workflow.

The sociodemographic and medical/family history features were selected from each participant’s medical and sociodemographic profile. These features were age, gender and years of education, maternal and paternal family history of AD, number of copies of the APoE ε4 alleles (abbreviated as APoE4). Reprocessed using 1-hot encoding, the APoE4 feature can take a value of 0, 1 or 2, representing the number of copies of the APoE4 allele. The CFA scores were collated from Alzheimer’s Disease Assessment Scale (ADAS), Cognitive Battery Assessment, Clinical Dementia Rating (CDR), Mini-Mental State Exam (MMSE), Modified Hachinski Ischemia Scale, Neuropsychological Battery Test, logical memory immediate recall test (LMIT), logical memory delayed recall test (LMDT), the Neuropsychological Inventory (NPI) and the Geriatric Depression Scale (GDS). As well as total scores for all these assessments, individual question scores from ADAS and individual subscales from NPI were included in the dataset. Other CFAs and individual

CFA subscales from ADNI were not included due to the large amounts of missing data for the set of participants who had undergone PET-MRI scans. For tau PET neuroimaging data, the [^18^F]AV-1451 tracer for detecting tau deposition was used [14]. After data merging and pre-processing, the dataset comprised 559 samples representing 363 cognitively normal (CN) individuals, 137 MCI (mild cognitive impairment) individuals and 59 Alzheimer’s disease (AD) participants.

### 2.2 Data Preparation

Prior to data pre-processing, sociodemographic, medical/family history and CFAs were combined using each participant’s unique identifier (ID). Inevitably, some participants were not present for a portion of the assessments without MRI and PET brain scans, which resulted in about 6% missing values. Relatively simple and sound imputation techniques were adopted in favour of more technical approaches as they tend to provide competitive performance with the absence of the computational and technical complexity Specifically, rows that contained sporadic missing values were imputed by mean imputation for numeric values and modal imputation for categorical values. For missing sociodemographic values a similarity matrix [16] was used for imputation, affecting 6% of the entire dataset in total. Approximately 10 members that showed close resemblance to the target row were provided by the resulting matrix, ultimately yielding a participant of greatest similarity to use as a reference for imputation. Once all missing data imputation had been completed, the dataset was normalised. Negative columns were initially isolated and normalised by increasing all values in the column by the absolute of the minimum value in that column.

### 2.3 Tau-PET and MRI Data Pre-processing

The pre-processing steps for the PET and MRI data are outlined as follows. Each PET scan was co-registered with its associated MRI scan before normalising to a predetermined AD template, whereby the resulting images were corrected for partial volume estimation (PVE) using the SPM toolbox PETPVE12 [17]. This reference serves as a template for dictating measures from the MRI scans. The values were sampled from brain regions defined by the Desikan-Killiany atlas [18] where the resulting labels were formatted using this custom template. Ultimately, this process yielded 177 neuroimaging features per participant based on their combined PET-MRI data. These were combined with the other data features and all features were called using min-max normalisation.

### 2.4 Unsupervised Learning, Feature Selection and Class Re-labelling

After data normalisation (see Section 2.2), the dataset of 224 features and 559 samples was subjected to dimension reduction using the uniform manifold approximation and project (UMAP) for dimension reduction and data visualisation in lower dimensional space [19]. Unsupervised manifold learning allows for efficient embedding of nonlinear data points while maintaining the relative distance or local connectivity of those points with respect to one another. For this study, a UMAP clustering was implemented with a large nearest neighbour parameter (30 neighbours) to avoid focusing on very local structures — a minimal distance value (0) was set to improve cluster density. 5 dimensions of the UMAP space was selected, based on visual inspection of the compactness of AD diagnosis cases distributed along these dimensions (see Results). Then, 3-dimensional UMAP plots were presented for data visualisation and clustering purposes. Each data point in each UMAP cluster was first assigned its original label (CN, MCI, or AD) based on clinician diagnosis before having some of the data points re-labelled (see below). Next, we used the unsupervised learning, *k*-means clustering [20], to identify discrete clusters within the UMAP space. *k*-means clustering had also been applied successfully on AD datasets in previous studies [21, 22] including the ADNI dataset. Additionally, feature selection by the information gain algorithm [23] was implemented with the FSelector package in R [24] to identify the top 10 features most associated with membership of a particular cluster for the originally labelled data and later, the re-labelled data. Once the key feature characteristics of each cluster were identified, the clusters were each given an appropriate unique name.

One of the clusters based on UMAP and *k*-means processing was subsequently identified to uniquely consist of a majority of clinically diagnosed AD cases; hence named the “AD” cluster. However, there were a few clinically diagnosed CN and MCI cases in this cluster, and some clinically diagnosed AD cases outside of the cluster. We re-labelled the CN and MCI cases in Cluster AD as AD cases. These relabelled cases were validated post-hoc based on their tau-PET and Aβ levels, which were found to be similar, while being intermediate between the non-re-labelled AD and non-AD cases. The re-labelled data was used for training the GNN model, which will be compared to GNN trained using the original clinician diagnosis labels (see Section 2.5).

### 2.5 Graph Neural Network (GNN) for Classification

The robust meta-learning-based auto-metric graph neural network (AMGNN) classifier developed by [12] was used to classify the data using separately, the original clinician diagnostic class labels and the new class labels. As in [12], a few features of the data were used to create the graph for the neural network classifier, with the rest of the data features processed in the context of the graph relationships. In this work, gender, age, education, and family history data were used to build the graph for the classifier. The node classification of this small graph was done by randomly selecting samples from the training dataset as a meta-task to train the AMGNN. Based on several meta-task training runs, the AMGNN model can then be used to classify unknown label nodes in a new graph [12]. Overall, the network consisted of 2 GNN layers, alongside a total of 4 CNN layers. For further details, please refer to [12] and https://github.com/mac-n/Clustering-GNN.

The dataset was split into training and testing datasets, at 80% and 20%, respectively. The model was trained over 300 iterations using data batches of 64 rows before updating the loss parameter. Model performance was tested using one-shot learning which included a single training sample in each batch [25]. This was repeated for both datasets; one with the original clinical diagnosis labels and the other based on cluster-based re-labelling. Balanced accuracy and multiclass area-under-the-curve (AUC) (calculated by averaging onevs-all AUC for each class) were visualized throughout training and testing process using TensorBoard, a visualization framework. Finally, the experiment was repeated 30 times to check for robustness and variability in performance. The rate of convergence during training sessions was also assessed.

### 2.6 Data and Code Availability

Python and R codes for clustering, data analysis and classification can be found in the code repository provided in the link https://github.com/mac-n/Clustering-GNN which includes the Python environment set up, dataset visualization, and some of the dataset preparation procedures and modelling process. The original ADNI dataset was not included as part of this repository. However, the final processed (and anonymized) dataset used for GNN model development has been provided for GNN model demonstration. Requests to access the original datasets should be directed to ADNI (http://adni.loni.usc.edu/).

## 3 Results

### 3.1 Distinctive Data Cluster Characteristics in Low Dimensions

Using the dimensional reduction UMAP method (see Section 2.4), the initial dataset of 224 features were projected to 5 UMAP dimensions (Figure 2A). In the figure, the classes CN (dark purple), MCI (light purple) and AD (yellow) labelled by clinician diagnosis are denoted in different colours. A 5-dimensional UMAP space was selected, as visual inspection of the AD diagnosis information as distributed along the 5 dimensions demonstrated that the AD cases were clustered tightly together along some of these dimensions (Figure 2A, yellow points). A simpler 3dimensional UMAP projection is illustrated in Figure 2B. Using *k*-means clustering, it can be observed that there were 5 distinct clusters (denoted by different colours). If we indicated the data points with clinical diagnosis (CN, MCI, or AD), we found that the distribution of clinical diagnosis did not generally conform well to the 5 discrete clusters (Table 1).

**Figure 2.**
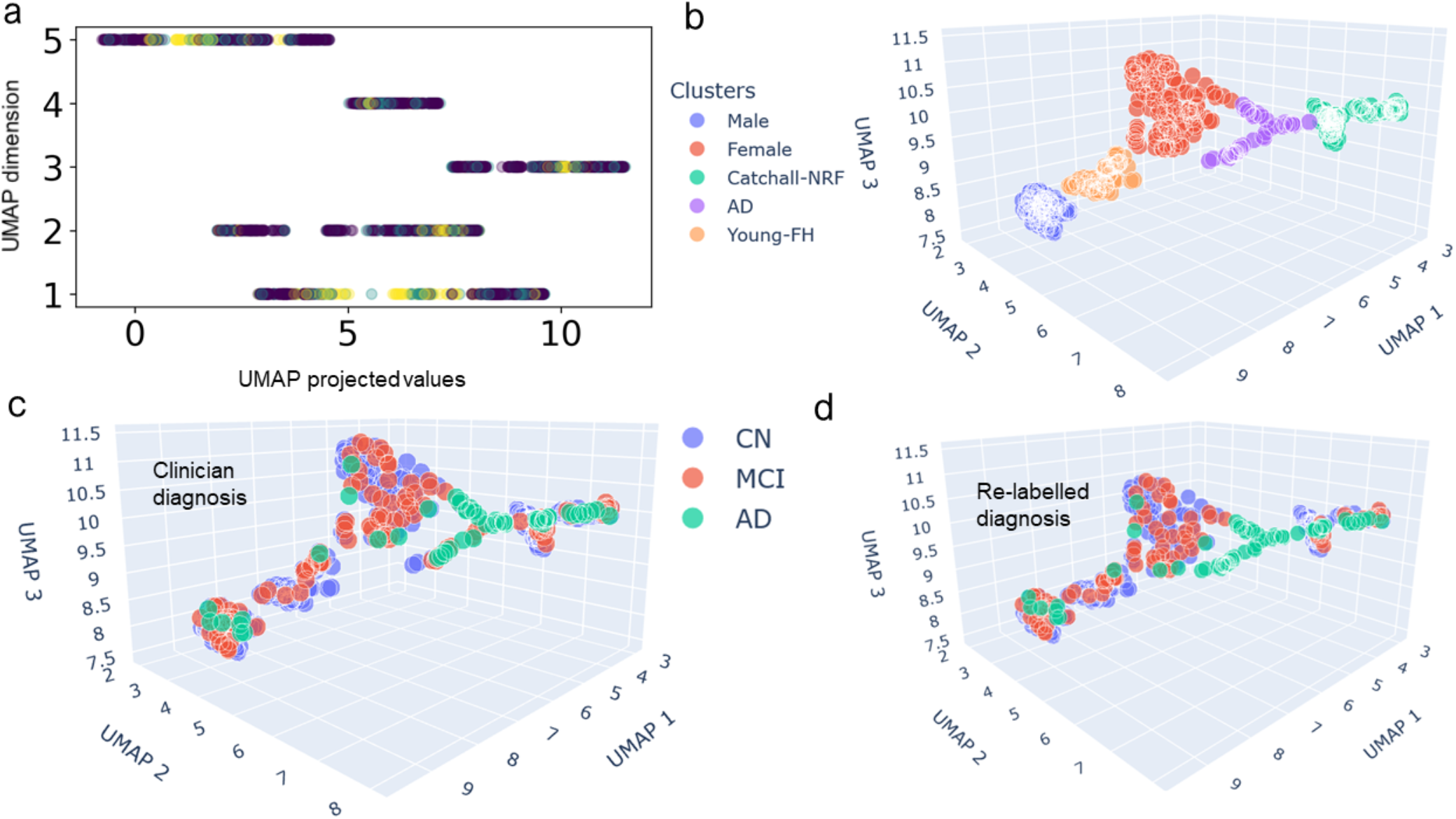
Data clusters in UMAP spaces. (a) Original data projected onto UMAP’s 5 dimensions with distribution of projected values. Colour labelling of classes (clinician diagnosis): CN (dark purple), MCI (light purple) and AD (yellow). **(b)** Data clusters in 3-dimensional UMAP space. Different colours to indicate the distinct clusters determined by k-means clustering information gain and tau PET imaging features. Cluster names were based on the unique feature characteristics of each cluster. (c-d) 3-dimensional UMAP with original clinician diagnostic labels (c) and with cluster-based class re-labelling (d). Colour labelling for (d) as in (c).

**Table 1.** Clusters’ sociodemographics and clinician diagnosis.

| Cluster Label | No. of Cases | Age (Years) | Gender: M/F (%) | Education (Years) | Clinician Diagnosis: CN/MCI/AD (%) |
| --- | --- | --- | --- | --- | --- |
| AD | 50 | 75.1 ± 9.8 | 30/20 (60/40) | 16.7 ± 2.5 | 6/18/26 (12/36/52) |
| Catchall | 166 | 74.6 ± 7.2 | 70/96 (42/58) | 16.3 ± 2.6 | **114/32/20 (69/19/12)** |
| Female | 131 | 75.9 ± 8.2 | 0/**131** (0/100) | 16.0 ± 2.4 | **96/28/7 (73/21/5)** |
| Male | 131 | **78.8** ± 6.9 | **131/0** (100/0) | 17.1 ± 2.5 | 82/44/5 (63/34/4) |
| Young-FH | 81 | **71.3** ± 6.4 | 35/46 (43/57) | 17.1 ± 2.1 | **65/15/1 (80/19/1)** |

Table 1 shows a summary of the sociodemographic and distribution of clinician diagnosis in each of the 5 clusters. In terms of years of education, all the clusters had similar average values around 16-17 years. The 2 most distinctive clusters were the gender specific clusters in which only male or female cases exist. This was consistent with previous unsupervised learning studies using different data [22, 26]. Hence, we named these two clusters as Male and Female (Table 1). Interestingly, the Male cluster had the oldest average age among all clusters and had a higher proportion of MCI cases than the Female cluster. In comparison, there was a cluster with the youngest average age (71 years old) with about equal proportion of male and female participants, and which also had the highest proportion of CN cases. For now, we named this cluster the Young cluster. Another distinctive cluster is one with the largest proportion of AD cases (52%), despite not having the oldest average age. We named this cluster as the AD cluster even though MCI cases constituted 36%. The final clusters consisted of almost equally mixed gender and with a substantially high number of CN cases. For now, we named this the Catchall cluster.

We next analysed the clusters’ characteristics using a feature selection method – information gain of the data features with respect to cluster labels (Section 2.4), with the hope that this method would shed light on the Catchall cluster. The top ranked features found by information gain that could distinguish the clusters were (ranked from the most important feature): APoE4, gender, (history of) mother with AD, ctx.rh.inferiorparietal, ctx.lh.middletemporal, lh. Amygdala, ctx.lh.inferiorparietal, ctx.lh.lateraloccipital, wm.lh.entorhinal, and wm.lh.inferiortemporal. APoE4, gender and (history) of mother with AD were identified as the top 3 features. Moreover, the list was dominated by various tau PET-MRI imaging features (7 out of 10 features). Hence, we analysed the clusters using the ctx.lh.inferiorparietal (left inferior parietal cortex) and lh.Amygdala (left amygdala) features, based on their suggested links to early stages of MCI and AD [27, 28].

Table 2 summarises the statistics of the values of ctx.lh.inferiorparietal and lh.Amygdala features across the 5 clusters. We can see that Cluster AD consistently had the largest values for both the statistics regardless for both brain regions (Table 1, bold text), as was expected. Interestingly, Cluster Catchall had the second largest values for most of the statistics (Table 1, bold text). These could be due to the 12% of clinically diagnosed AD cases (second highest cases among all the clusters), albeit 69% CN and 19% MCI cases (Table 1). Hence, this cluster might consist of early MCI or AD stage, with or without formal diagnosis by clinicians – i.e. with potential neurological risk factor (NRF). We therefore renamed this cluster as Cluster Catchall-NRF. This was validated by comparing participant CSF Aβ level, which was not used in the clustering, across the clusters. Table 2 shows that Cluster AD has the highest level of amyloid pathology (lowest value) and Cluster Catchall-NRF has the second highest level (second lowest value). Note that CSF Aβ levels were not available for Cluster Young-FH due to a large number of missing values.

**Table 2.** Summary of the statistics of participants’ tau PET-MRI neuroimaging features, ctx.inferiorparietal and lh.Amygdala, with respect to the 5 clusters, and participant CSF Aβ levels, a variable not used in the clustering model.

| Cluster | ctx.inferiorparietal | lh.Amygdala | CSF Aβ |
| --- | --- | --- | --- |
| AD | **2.363 ± 1.277** | **1.831 ± 0.439** | **637±257** |
| Catchall-NRF | **1.506 ± 0.362** | **1.336 ± 0.342** | **1038±436** |
| Female | 1.438 ± 0.212 | 1.185 ± 0.185 | 1346±372 |
| Male | 1.319 ± 0.164 | 1.198 ± 0.16 | 1260±423 |
| Young-FH | 1.395 ± 0.16 | 1.196 ± 0.165 | n/a |

### 3.2 Re-labelling of Classes and Validation

When clinician diagnosis (CN, MCI or AD) was indicated in these clusters (Figure 2C), we observed overlaps between the UMAP clusters and the clinician diagnostic labelled classes, with substantial mixing between the classes in some of the clusters. As mentioned earlier, one of the clusters (Cluster AD) particularly had substantially purer (AD) cases. Hence, we hypothesised that we could re-label the CN and MCI cases in Cluster AD as AD cases (CN/MCI-to-AD). The re-labelled data points are visually shown in Figure 2D (compared to Figure 2C). The kappa index of agreement between original clinician diagnosis and re-labelled diagnosis is 0.917.

Next, based on the selected features we made use of CFA feature PHC_MEM, neuroimaging features lh.Amygdala and and wm.lh.entorhinal to perform a post-hoc check for the relabelled cases, given the known link of the amygdala to early-stage AD [28-30]. Table 3 shows the mean and standard deviation for these 2 features for the 3 cases: (i) AD-to-MCI outside Cluster AD; (ii) remained AD cases in Cluster AD; and (iii) remained non-AD cases outside Cluster AD. It can be observed that for the CN/MCI-to-AD (in Cluster AD) relabelled cases, the values for both the lh.Amygdala and wm.lh.entorhinal features’ values were intermediate between the values of the remained AD cases and remained non-AD cases. The level of CSF Aβ [15] for each group, a variable not used in the clustering, is also shown in Table 3. Interestingly, it can be seen that CSF Aβ pathology was more advanced (lower value) in the re-labelled cases than in the clinically diagnosed AD cases (t-test: p=0.007), even though the mean age of the clinically diagnosed AD cases was older (p=0.08). Therefore, the diagnostic re-labelling was well supported by the tau-PET neuromarkers and the CSF Aβ marker. We did not re-label any cases which were diagnosed as AD by clinicians but not by our algorithm (i.e. there is no AD-to-CN or AD-to-MCI). There were 33 such cases in the data, but there appeared to be little clinical justification for re-labelling them; they are characterised by slightly lower tau-PET levels and older age than other AD cases.

**Table 3.** Mean age (in years) with mean and standard deviation of PHC_MEM (CFA) and lh.Amygdala and wm.lh.entorhinal (neuroimaging), and Aβ (CSF) features with respect to the re-labelled CN/MCI-to-AD cases, compared to non-AD cases and non-re-labelled AD cases. The cases diagnosed with AD which fell outside the AD cluster are shown at the bottom (below dashed-dotted line), though these cases were not re-labelled. CSF Aβ feature was not used in the clustering.

| Types of Cases | No. of Cases | Age | PHC_MEM | lh. amygdala | wm.lh. entorhinal | CSF Aβ |
| --- | --- | --- | --- | --- | --- | --- |
| Non-AD | 476 | 75.3 | $0.71 \pm 0.60$ | $1.21 \pm 0.19$ | $1.23 \pm 0.22$ | 1257 |
| CN/MCI to AD | 18 | 74.2 | $0.49 \pm 0.11$ | $1.67 \pm 0.39$ | $1.71 \pm 0.34$ | 621 |
| Remained AD | 59 | 77.9 | $0.31 \pm 0.11$ | $1.84 \pm 0.44$ | $1.94 \pm 0.51$ | 851 |
| AD outside AD cluster | 33 | 79.3 | $0.34 \pm 0.12$ | $1.74 \pm 0.50$ | $1.81 \pm 0.50$ | 845 |

### 3.3 More Accurate Classification for cluster-based labels

With the re-labelled data, we can now train the GNN model for 3-class (CN, MCI, AD) classification by [12] (Sections 2.5-2.6) using both the originally labelled data based on clinician diagnosis and the re-labelled data (Section 3.2). The convergence trends for the first 6 repetitions of each experiment are shown in Figure 3. Over the 30 repetitions, the GNN model using the re-labelled data achieved an average multiclass AUC of 95.1±0.04%, which should be compared to the AUC using the originally labelled data of 91.7±0.07% accuracy. A t-test comparing the AUC values over the 30 repetitions yielded a p-value of 0.02. Balanced accuracy was also recorded: 93.6 ± 0.05% with the re-labelled data and 89.3±0.06% with the original labelled data.

**Figure 3.**
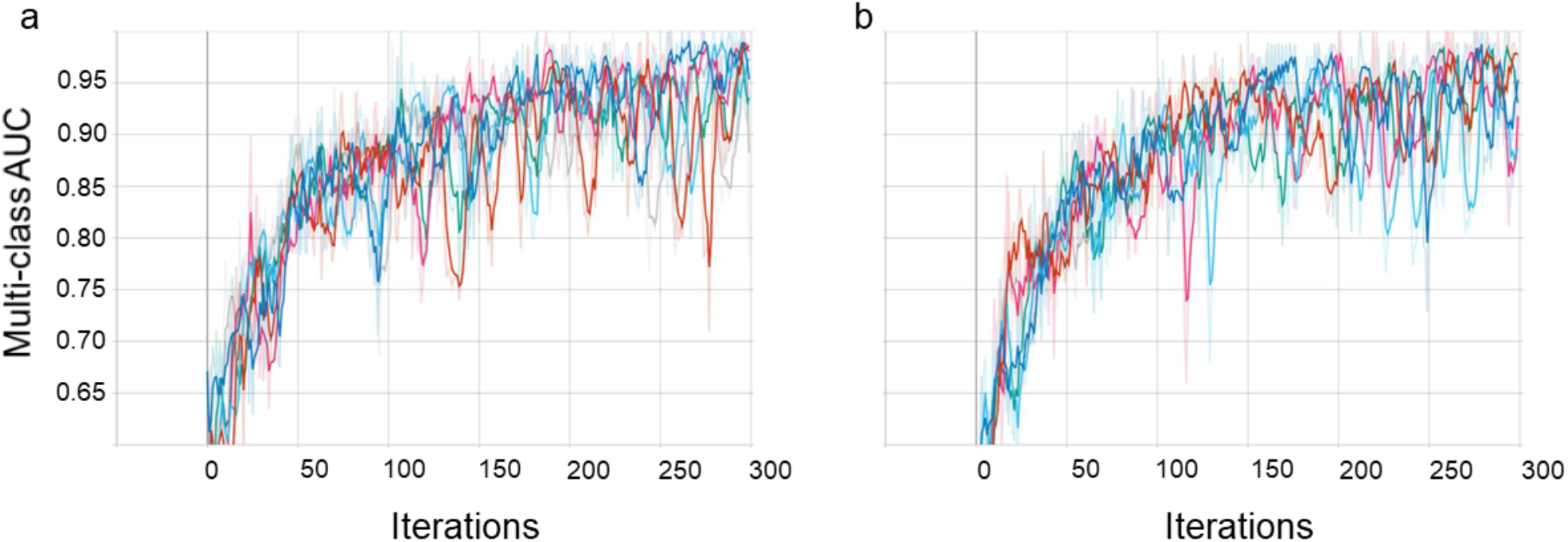
GNN (3-class) AUC across the first 6 runs. (a-b) Convergence for GNN model using originally labelled (a) and re-labelled (b) data.

## 4 Discussion

In this work, we have successfully incorporated both unsupervised learning (UMAP and *k*-means clustering) and supervised learning (information gain feature selection) to provide insights into heterogeneous AD data, and subsequently using supervised learning with GNN for diagnostic classification.

Prior to applying GNN for AD classification, we made use of nonlinear dimensional reduction UMAP (Figure 2) and projected the data into a 5-dimensional UMAP (Figures 1A-D) space for deeper insights and for guidance in the relabelling (Figure 2D). Five discrete data clusters were identified using *k*-means clustering (Figure 2B): a majority AD cluster, a fully male cluster, a fully female cluster, a cluster of younger participants with parental history of AD (“Young-FH” cluster), and a relatively unknown “Catchall” cluster. It is interesting to speculate that the Young-FH could be due to relatively younger participants who were concerned about their own health given that their parents had a history of AD. The 2 gender specific clusters identified seemed to be in line with previous studies [22, 26, 31].

The results of the clustering were additionally validated by data exogeneous to the clustering algorithm, specifically CSF Aβ, which is a biomarker characteristic of prodromal AD [32]. The cases which were classified into the AD cluster which did not have a clinical AD diagnosis were characterized by higher average levels of CSF Aβ than even diagnosed AD cases, at a younger age. Similarly, cases which were classified into the NRF cluster were characterized by higher Aβ levels than members of any other cluster apart from the AD cluster. In future work it could be interesting to explore the possibility that the algorithm is identifying a specific subtype of AD, since cases where the algorithm and the clinicians disagreed differed significantly on age and CSF Aβ levels.

Future work, given ongoing availability of longitudinal tau-PET data from ADNI, could use the same cluster mapping to investigate AD progression, validate the re-labelling, and the hypothesis that the NRF cluster is a high-risk group. Although unsupervised clustering for AD prognosis has yet to be performed on tau-PET data, there are examples of clustering on other markers successfully predicting AD progression [22, 26, 33, 34].

In terms of supervised learning, GNNs have been introduced into AD classification studies in a variety of ways using different types of data, especially MRI and PET brain data [6, 8-10, 35, 36]. More recent advancements of GNNs on AD have been proposed to provide more flexibility [11, 12]. However, these studies have not considered detailed tau PET neuroimaging data, a limitation given the closer alignment of tau PET with AD stages [13, 14] than e.g. amyloid PET, and the recent approval of its use by the U.S. FDA [37]. Importantly, these studies, together with the literature on unsupervised learning approaches applied to AD [5-7, 21, 22, 26, 31, 33, 38] have not used cluster-based class re-labelling for classification of AD. Here, we have made use of a robust auto-metric GNN model [12]. Importantly, we made use of UMAP-cluster based re-labelling to train the GNN model. The re-labelled data led to a more accurate GNN model for detecting CN, MCI and AD groups. Future work could explore deep learning classifiers combined with automatic feature extraction [39] which has the potential to provide deeper insights into the combinations of data features and brain regions most relevant to the disease classification [40].

Taken all together, the high diagnostic accuracy of the relabelling approach in this work highlights the potential for data-driven methods to be incorporated into the diagnostic process for AD. This study reinforces the value of methods such as unsupervised clustering method, to derive new patterns and sub-groups from existing datasets and enhance the current clinical methodology for AD diagnosis and risk stratification.

## Data Availability

All data produced in the present study are available upon reasonable request to the authors

## Acknowledgements

This work was supported by the European Union’s INTERREG VA Programme, managed by the Special EU Programmes Body (SEUPB; Centre for Personalised Medicine, IVA 5036). The views and opinions expressed in this paper do not necessarily reflect those of the European Commission or the Special EU Programmes Body (SEUPB). Data collection and sharing for this project was funded by the Alzheimer’s Disease Neuroimaging Initiative (ADNI) (National Institutes of Health Grant U01 AG024904) and DOD ADNI (Department of Defense award number W81XWH-12-2-0012). ADNI is funded by the National Institute on Aging, the National Institute of Biomedical Imaging and Bioengineering, and through generous contributions from the following: AbbVie, Alzheimer’s Association; Alzheimer’s Drug Discovery Foundation; Araclon Biotech; BioClinica, Inc.; Biogen; Bristol-Myers Squibb Company; CereSpir, Inc.; Cogstate; Eisai Inc.; Elan Pharmaceuticals, Inc.; Eli Lilly and Company; EuroImmun; F. Hoffmann-La Roche Ltd and its affiliated company Genentech, Inc.; Fujirebio; GE Healthcare; IXICO Ltd.; Janssen Alzheimer Immunotherapy Research & Development, LLC.; Johnson & Johnson Pharmaceutical Research & Development LLC.; Lumosity; Lundbeck; Merck & Co., Inc.; Meso Scale Diagnostics, LLC.; NeuroRx Research; Neurotrack Technologies; Novartis Pharmaceuticals Corporation; Pfizer Inc.; Piramal Imaging; Servier; Takeda Pharmaceutical Company; and Transition Therapeutics. The Canadian Institutes of Health Research is providing funds to support ADNI clinical sites in Canada. Private sector contributions are facilitated by the Foundation for the National Institutes of Health (www.fnih.org). The grantee organization is the Northern California Institute for Research and Education, and the study is coordinated by the Alzheimer’s Therapeutic Research Institute at the University of Southern California. ADNI data are disseminated by the Laboratory for Neuro Imaging at the University of Southern California.

